# Magnitude and Dynamics of the T-Cell Response to SARS-CoV-2 Infection at Both Individual and Population Levels

**DOI:** 10.1101/2020.07.31.20165647

**Authors:** Thomas M. Snyder, Rachel M. Gittelman, Mark Klinger, Damon H. May, Edward J. Osborne, Ruth Taniguchi, H. Jabran Zahid, Ian M. Kaplan, Jennifer N. Dines, Matthew T. Noakes, Ravi Pandya, Xiaoyu Chen, Summer Elasady, Emily Svejnoha, Peter Ebert, Mitchell W. Pesesky, Patricia De Almeida, Hope O’Donnell, Quinn DeGottardi, Gladys Keitany, Jennifer Lu, Allen Vong, Rebecca Elyanow, Paul Fields, Julia Greissl, Lance Baldo, Simona Semprini, Claudio Cerchione, Fabio Nicolini, Massimiliano Mazza, Ottavia M. Delmonte, Kerry Dobbs, Rocio Laguna-Goya, Gonzalo Carreño-Tarragona, Santiago Barrio, Luisa Imberti, Alessandra Sottini, Eugenia Quiros-Roldan, Camillo Rossi, Andrea Biondi, Laura Rachele Bettini, Mariella D’Angio, Paolo Bonfanti, Miranda F. Tompkins, Camille Alba, Clifton Dalgard, Vittorio Sambri, Giovanni Martinelli, Jason D. Goldman, James R. Heath, Helen C. Su, Luigi D. Notarangelo, Estela Paz-Artal, Joaquin Martinez-Lopez, Jonathan M. Carlson, Harlan S. Robins

## Abstract

T cells are involved in the early identification and clearance of viral infections and also support the development of antibodies by B cells. This central role for T cells makes them a desirable target for assessing the immune response to SARS-CoV-2 infection. Here, we combined two high-throughput immune profiling methods to create a quantitative picture of the T-cell response to SARS-CoV-2. First, at the individual level, we deeply characterized 3 acutely infected and 58 recovered COVID-19 subjects by experimentally mapping their CD8 T-cell response through antigen stimulation to 545 Human Leukocyte Antigen (HLA) class I presented viral peptides (class II data in a forthcoming study). Then, at the population level, we performed T-cell repertoire sequencing on 1,815 samples (from 1,521 COVID-19 subjects) as well as 3,500 controls to identify shared “public” T-cell receptors (TCRs) associated with SARS-CoV-2 infection from both CD8 and CD4 T cells. Collectively, our data reveal that CD8 T-cell responses are often driven by a few immunodominant, HLA-restricted epitopes. As expected, the T-cell response to SARS-CoV-2 peaks about one to two weeks after infection and is detectable for at least several months after recovery. As an application of these data, we trained a classifier to diagnose SARSCoV-2 infection based solely on TCR sequencing from blood samples, and observed, at 99.8% specificity, high early sensitivity soon after diagnosis (Day 3–7 = 85.1% [95% CI = 79.9-89.7]; Day 8–14 = 94.8% [90.7-98.4]) as well as lasting sensitivity after recovery (Day 29+/convalescent = 95.4% [92.1-98.3]). These results demonstrate an approach to reliably assess the adaptive immune response both soon after viral antigenic exposure (before antibodies are typically detectable) as well as at later time points. This blood-based molecular approach to characterizing the cellular immune response has applications in clinical diagnostics as well as in vaccine development and monitoring.

## Introduction

The adaptive immune response to infection includes both a cellular and humoral component. The cellular immune response is mediated by T cells, which play a role in direct killing of virus-infected cells via cytotoxic (CD8) T cells as well as helping to direct the overall immune response through helper (CD4) T cells. The humoral immune response also includes CD4 T cells which assist B cells to differentiate into plasma cells and subsequently produce antibodies specific to a targeted antigen. As T cells are involved in the early identification and clearance of viral infections by both cellular and humoral immunity, they are a desirable target for assessing SARS-CoV-2 exposure (Grifoni 2020, Weiskopf 2020, Peng 2020, Sekine 2020, Altmann 2020).

Healthy adults have ~10^12^ circulating T cells expressing approximately 10^7^ unique TCRs (Robins 2009). This diversity allows the full repertoire of T cells to potentially recognize a wide variety of peptide antigens displayed by HLA molecules on the surface of cells. When a naïve T cell is activated in response to recognition of a cognate antigen presented by a specialized antigen presenting cell, it undergoes clonal expansion, resulting in an exponentially increasing number of genetically identical T cells. Due to the extreme sequence diversity possible among TCR rearrangements, particularly the TCR-beta chain, each observed TCR sequence is essentially a unique tag for a clonal lineage of T cells. Thus, the number of copies of each TCR sequence represents the number of T cells in that clonal lineage and provides information about the natural history of T-cell clonal expansions. Measuring the cellular immune response can provide a view into the state of the overall immune response, and several qualities of the adaptive cellular immune response suggest a T-cell-based assay may fulfill unmet clinical needs. In general, the T-cell immune response is: 1) Sensitive: T cells detect even a very small amount of antigen; 2) Specific: TCRs bind only to specific antigens; 3) Naturally amplified: T cells proliferate and clonally expand upon recognition of small quantities of specific antigen via their TCRs; 4) Systemic: T-cell clones circulate throughout the body in the blood; and 5) Persistent: a subset of T cells are maintained following clonal contraction in long term memory (Robins, 2013, DeWitt 2015, Dash 2017, Glanville 2017, DeWitt 2018). The T-cell response is typically the first component of the adaptive immune response that can be measured, within days from initial pathogen exposure, and after clonal expansion and transition into memory can persist for years even when antibodies become undetectable. In the context of coronavirus infections, persistent T cells specific for SARS-CoV-1 have been routinely detected in studies in the years following the initial SARS outbreak (Peng 2006, Tang 2011), including at least a decade after initial infection (Ng 2016). Subjects show lasting memory T-cell populations to SARS-CoV-1 even as IgG antibodies and peripheral memory B cells become undetectable in a majority of convalescent subjects (Tang 2011). Similarly, T cells responsive to the Middle East respiratory syndrome (MERS) coronavirus were observed in the absence of detectable antibodies (Zhao 2017).

Standard methods to assess the cellular immune response to a pathogen are based on T-cell recognition of target antigens. Conventional immune monitoring assays, including ELISpot and ICS, rely on functional T-cell responses and require live T cells, thus limiting standardization and throughput. The emergence of the COVID-19 pandemic has generated the urgent need for a scalable molecular assay to assess the T-cell response to SARS-CoV-2. In response, Adaptive Biotechnologies and Microsoft have applied previously developed platforms to create immunoSEQ^®^ T-MAP™ COVID, a TCR sequence-based approach to quantitatively assess the T-cell response to SARS-CoV-2. This approach utilizes a multiplexed experimental platform to interrogate T-cell repertoires with large numbers of query antigens to identify SARS-CoV-2-specific TCRs in the context of HLA (Klinger 2015). We have deeply characterized 61 COVID-19 subject samples against 545 potential peptide antigens to profile the CD8 immune response. We have further sequenced 1,815 blood samples from 1,521 COVID-19 cases with immunoSEQ^®^ in order to identify a robust set of SARS-CoV-2 specific CD4 and CD8 TCRs from a fixed number of blood cells (Carlson 2013, Robins 2012). All of these data are available as part of the public ImmuneCODE data release at https://clients.adaptivebiotech.com/pub/covid-2020 (Nolan 2020).

Taken together, these approaches allow the development of a map between TCR sequences and SARSCoV-2 specific antigens, as well as the identification of public SARS-CoV-2 specific TCRs shared across individuals. This approach allows us to characterize many of the antigens involved in a T-cell immune response. We also capture a measure of the clonal breadth (the estimated proportion of distinct T-cell clonal lineages in a repertoire that are SARS-CoV-2 specific) and depth (related to the relative frequency of SARS-CoV-2-specific T-cell clones in a repertoire), as well as the dynamics of the cellular immune response to a SARS-CoV-2 infection over time. The exact antigens targeted are elucidated for several of these clones, which may allow for mapping a vaccine response in comparison to the response in a natural infection (DeWitt 2015). Moreover, a collection of public SARS-CoV-2 TCRs form a robust diagnostic for recent or past infection of SARS-CoV-2. We report initial findings that the T-cell response is durable for at least 3 months after infection, which is the current limit of the samples available to assess.

## Results

### Identification of SARS-CoV-2-specific TCRs from COVID-19 subjects

To directly characterize the CD8 T-cell response to SARS-CoV-2, we applied MIRA (<u>M</u>ultiplex Identification of T-cell Receptor Antigen Specificity), which maps TCRs to antigens at high scale and specificity (Klinger 2015). 545 query peptides derived from across the SARS-CoV-2 genome were selected from HLA-I NetMHCpan predictions across multiple representative HLA types (Andreatta 2016, Nielsen 2003). These peptides were synthesized and assigned either individually or as groups of related peptides to one of 269 unique MIRA pools or “addresses” as described in the methods.

MIRA was performed on T cells derived from PBMCs collected from 3 acutely infected and 58 convalescent COVID-19 subjects. Overall, 23,179 unique SARS-CoV-2 specific CD8 TCRs were identified 25,442 times across all experiments. The identified TCRs mapped to 260 of the 269 pools, representing antigens from across the viral proteome (Figure 1a,b). Strong immune responses (assessed by total number of TCRs) as well as common immune responses (assessed by number of subjects with response to an antigen) were observed across the viral proteome.

**Figure 1:**
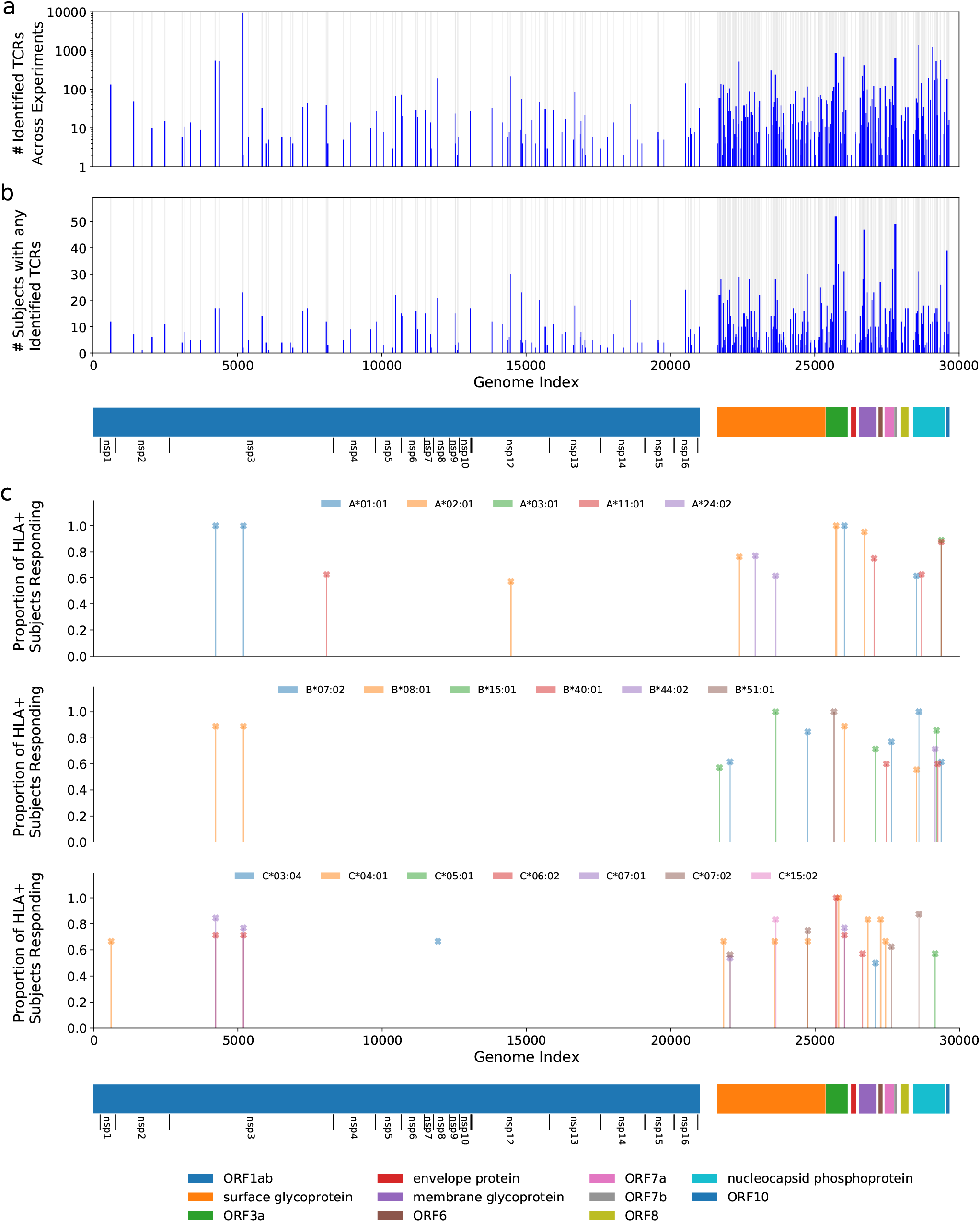
Magnitude and immunodominance of T-cell response to hundreds of potential SARS-CoV-2 antigens. Panel (a) shows the count of identified TCRs across experiments at each antigen position in the viral genome, and Panel (b) shows a similar representation for the count of subjects that had at least 1 TCR identified in the data at that antigen position. The blue bars represent these counts while the gray background indicates the areas covered by the tested antigens. Panel (c) shows the proportion of individuals with a given HLA that respond to a given antigen, restricting to immunodominant antigens. For this figure, we define *response* to mean a MIRA experiment using a subject who expresses the given HLA and for which the number of identified TCRs are more than 2-fold higher than the median number observed for experiments with donors who do not express the HLA. Only HLAs that are observed in at least ≥5 donors are considered, and only HLA-antigen pairs with at least 50% response rates and significant median-fold enrichment are shown. Note that no correction was made for HLA linkage disequilibrium. Detailed data and significance tests are available in Table S2. The eleven open reading frames from the virus are indicated below the plots including extra notation for the sixteen nonstructural proteins (nsp) encoded by ORF1ab.

We then explored the diversity of TCRs identified by MIRA across all the subjects by protein and by antigen. Figure 2a shows a clustergram of the protein-level response by subject, normalized to show the fraction of total TCRs identified per target. Figure 2b shows a similar analysis at the antigen-level, showing the 50 antigen locations with the most total TCRs observed across all subjects. A complete representation of the TCRs by antigen location is given in Supporting Table S1. Preliminary analyses indicate these response data were heavily skewed by antigen, with 70% of all TCR mappings accounted for by 14 antigen pools (Supporting Table S1). Similarly, responses to 8 antigens were observed in over half of the COVID-19 subjects’ MIRAs, suggesting these epitopes are frequently targeted during natural infection (Table S1).

**Figure 2:**
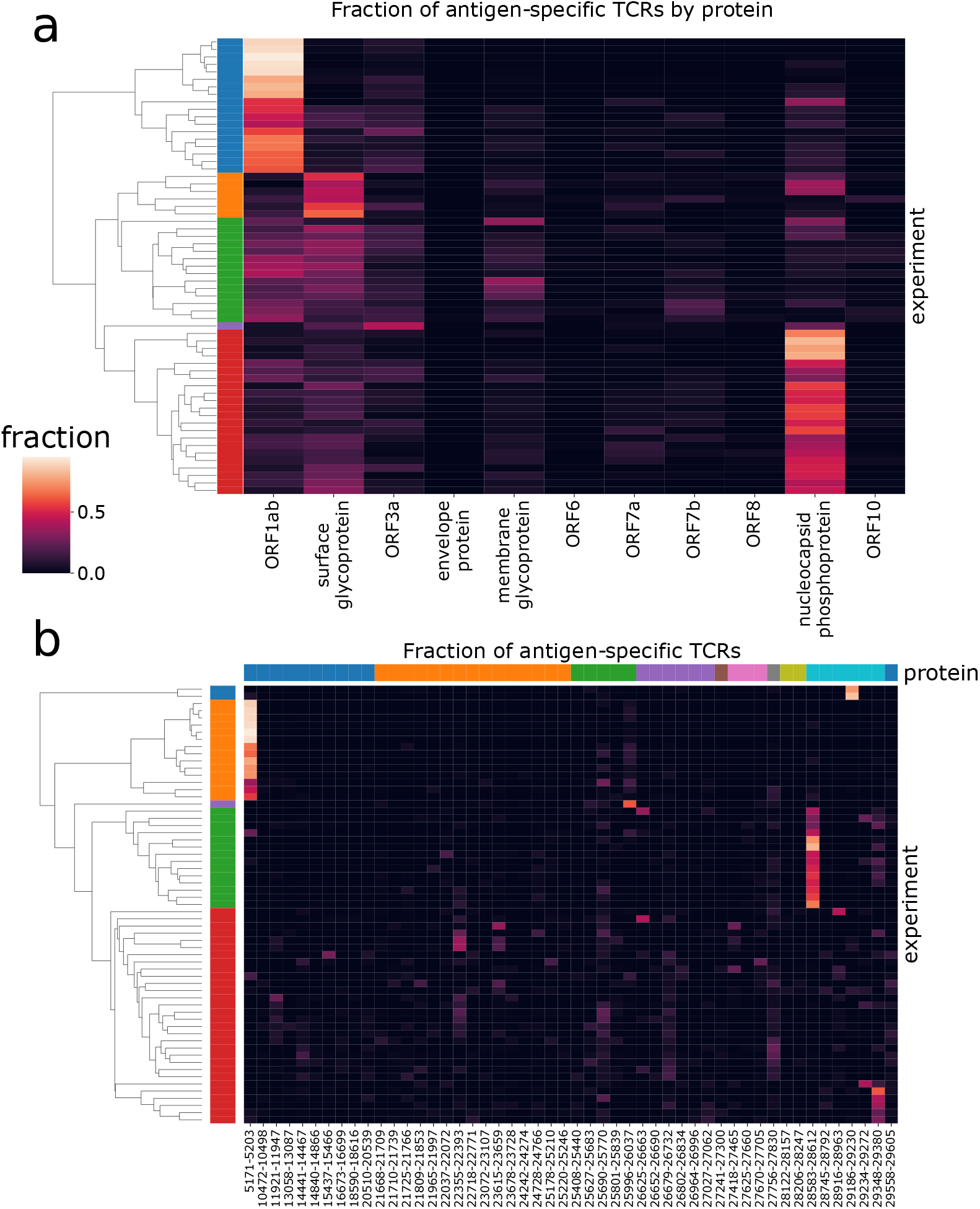
Patterns of antigen-TCR reactivity reveal immunodominance of some antigens. These two panels show clustergram plots of the (a) protein-level and (b) antigen-level signals across subjects. Each of the rows in the plot represents a distinct subject from the MIRA experiments; the left side label shows coloration for the top five subject clusters. In panel (a), the columns represent the 11 viral proteins in viral genome order. In panel (b), the columns represent the 50 antigens having the most donors with 1 or more TCRs reacting to them. They are sorted in viral genome order and colored by protein at the top. (Note that clustering is done independently for each panel to show the five farthest-separated clusters, and subject sets will vary by color across panels.)

Our results suggest that in many subjects the immune response is dominated by a large number of distinct T cells against just a few epitopes, which may result from distinct HLA presentation. Figure 2a shows about 30% of subjects (first cluster, blue) have a predominantly ORF1ab-directed response in terms of total distinct T-cell clones, which is primarily explained by the single peptide HTTDPSFLGRY. Similarly, about 35% of subjects (fifth cluster, red) have a predominantly nucleocapsid phosphoprotein response, represented by at least two dominant antigen positions. Another cluster (third cluster, green) shows a more distributed response across multiple proteins/antigens while the second cluster in orange has stronger surface glycoprotein response.

An HLA association analysis to the identified antigen-level clusters (Figure 2b) was performed using Fisher’s exact test. The second cluster (orange) is primarily explained by TCRs associated with ORF1ab:5171-5203 (single peptide HTTDPSFLGRY). There are 12 subjects in this cluster with HLA typing available; all 12 have HLA-A*01:01 demonstrating significant enrichment (p=2e-10) for this allele considering only 13 subjects have this allele in this dataset. This peptide is predicted to be presented by HLA-A*01:01 by NetMHCpan. Similarly, the fourth cluster (green) contains 11 cases with HLA typing and all 11 subjects (out of 13 in this dataset) have HLA-B*07:02 (p=4e-9). There are two overlapping peptides in this address (LSPRWYFYY and SPRWYFYYL); the latter is predicted to be presented by HLA-B*07:02 by NetMHCpan. Beyond this cluster-focused analysis, putative HLA restriction has been attributed to each of these pools using a Mann-Whitney’s U test over the number of mapped TCRs per experiment (Supporting Table S2), identifying 41 strong associations between antigens and HLA alleles. For 18 alleles, we identified at least one putative immunodominant epitope, which we defined as an HLA-antigen pair for which at least 50% of individuals with that allele respond to the antigen (see Figure 1c for details and definitions). These results are consistent with other recent reports of strong HLA-dependent CD8 T-cell responses to specific antigens (Nelde 2020, Ferretti 2020). These assignments and emerging immunodominance hierarchies will be further explored in later work as we continue to perform MIRA on cases and controls.

Overall, these results suggest that the basis of an individual immune response is both heterogeneous and influenced by HLA background; some subjects show large responses to just a few antigens from SARS-CoV-2 while others show a broader response. This analysis also identifies a short-list of highly immunogenic antigens to focus on for further characterization of the CD8 T-cell response across individuals. We are generating more data to extend these results, as well as adding in class II restricted antigens to further profile COVID-19 subjects’ CD4 T-cell responses.

### Identifying shared SARS-CoV-2-associated TCRs across the population

While the diversity of TCR recombination means that most TCR responses are “private” and will be infrequently seen in other individuals, a part of the T-cell response to a disease is “public” with the same amino acid sequences observed in many individuals, particularly in shared HLA backgrounds (Venturi 2008). Such disease-associated TCRs can be identified using a case/control design, as previously described for cytomegalovirus (Emerson 2017).

To this end, a dataset of 1,015 samples from individuals currently or previously infected with SARS-CoV2 were collected as part of the ImmuneCODE project (Nolan 2020). Immunosequencing was performed to sample the TCR repertoires as described in the Methods. Additionally, 3,500 repertoires from our database processed prior to March 2020 were identified as controls (see Supporting Table S3 for cohort summaries; for this preliminary analysis, we used the data available in the version 002 ImmuneCODE release). A lower T-cell fraction (suggesting lymphopenia) was observed in a number of the COVID-19 cases compared to healthy immune repertoires consistent with prior reports (Cao 2020) (Supporting Figure S1). Public COVID-19 associated TCRs, which we call “enhanced sequences”, were then identified using Fisher’s exact test, as described in the methods.

As a pilot study, enhanced sequences were identified using two cohorts, DLS (from New York, USA) and NIH/NIAID (from Italy), comprising a total of 483 cases, with 1,798 pre-March 2020 controls. A total of 1,828 enhanced sequences were identified from this first dataset which collectively distinguish cases from controls (Figure 3a). To establish high confidence in the enhanced TCR sequences identified for SARS-CoV-2, sequence identification was also performed independently for each of these two cohorts. A total of 309 enhanced sequences from the earlier set of 1,828 were identified independently across both studies. This degree of overlap in distinct populations demonstrates the generality of the signal that has been discovered, while also pointing to the opportunity that additional data have to identify more SARS-CoV-2 associated sequences. Notably, these enhanced sequences were also substantially enriched in our other held-out cohorts in this initial dataset, which totaled 397 cases from three additional cohorts (ISB, H12O and BWNW) and 1,702 additional controls (Figure 3b).

**Figure 3:**
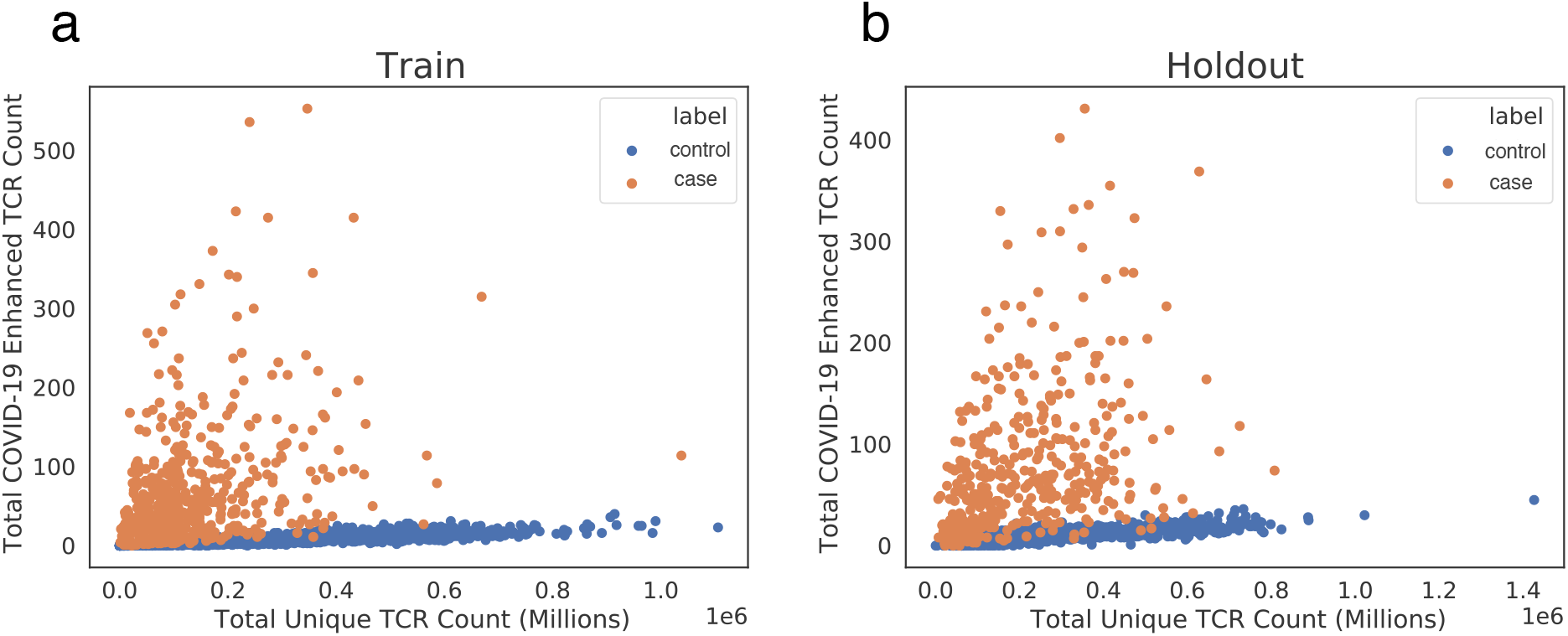
Public enhanced sequences associated with SARS-CoV-2 infection distinguish cases from controls. Panels (a) and (b) show the number of TCRβ DNA sequences in a subject that encode a SARS-CoV-2 enhanced sequence versus the total number of unique TCRβ DNA sequences sampled from that subject for a large number of cases and controls. Panel (a) represents the training set to identify this initial enhanced sequences list (DLS and NIH/NIAID cohorts), and panel (b) represents a hold-out set with no overlap with the training set (ISB, H12O and BWNW cohorts). Both panels show a similar number and separation of enhanced sequences in cases versus controls.

If these public associated enhanced sequences are SARS-CoV-2 specific, then a subset of them should overlap with the antigen-specific TCRs identified by the MIRA experiments. We identified a total of 368 exact matches to 59 different enhanced sequences from the set of 1,828 identified above. There were also 810 matches (from 394 distinct TCRs) to sequences that were only one amino acid change away (with identical V-gene and J-gene assignments) from 68 distinct enhanced sequences. Of the 59 different enhanced sequences with any exact matches, 36 (61%) were mapped to the HTTDPSFLGRY peptide from ORF1ab, with the remaining 23 mapping to 11 other antigen locations from across the proteome including two other ORF1ab addresses, four surface glycoprotein addresses, two nucleocapsid phosphoprotein addresses, and one each from ORF6, ORF10, and the envelope protein (see Supporting Table S1). Including near neighbors and other sequence-based clusters of TCRs would expand this count.

### Public disease-associated TCRs predict the breadth and depth of the antigen-specific T-cell response

To further explore the relationship between public disease-associated TCRs and largely private antigen-specific TCR datasets identified by MIRA, repertoire sequencing was performed on the COVID-19 subjects with MIRA data using the immunoSEQ assay. Although the current MIRA experiments are limited to CD8 T cells specific to the 545 HLA-I presented peptides in the MIRA panel, intersecting a subject’s MIRA-mapped TCRs with their immunoSEQ repertoire provides a lower bound estimate on the proportion of T cells in a subject that have likely expanded in response to SARS-CoV-2. Two specific quantities are of interest: the *clonal breadth* of the TCR repertoire, defined as the proportion of all unique TCR (DNA) clones that are SARS-CoV-2 specific; and the *clonal depth* of the TCR repertoire, related to the overall proportion of T cells that are SARS-CoV-2 specific (see Methods for precise definition).

Across 51 samples with paired immunosequencing and COVID-19 MIRA data, we observed a remarkable concordance between either the breadth (Figure 4a; Spearman rho = 0.62, p = 2e-6) or depth (Figure 4b; Spearman rho = 0.67, p = 6e-8) of an individual’s antigen-specific response as estimated by MIRA and that of the disease-specific response as estimated through public enhanced sequences. Notably, both clonal depth and breadth as measured by an individual’s MIRA response is typically an order of magnitude higher than that estimated by public clones, highlighting the extent to which MIRA is able to identify disease-associated TCRs, in addition to mapping TCRs to specific antigens. Nevertheless, for a small number of subjects, the clonal breadth and depth as estimated by public disease-specific clones is substantially higher than what is estimated by MIRA, likely indicating the role of CD4 T cells as well as CD8 T cells specific to antigens not included in the panel.

**Figure 4:**
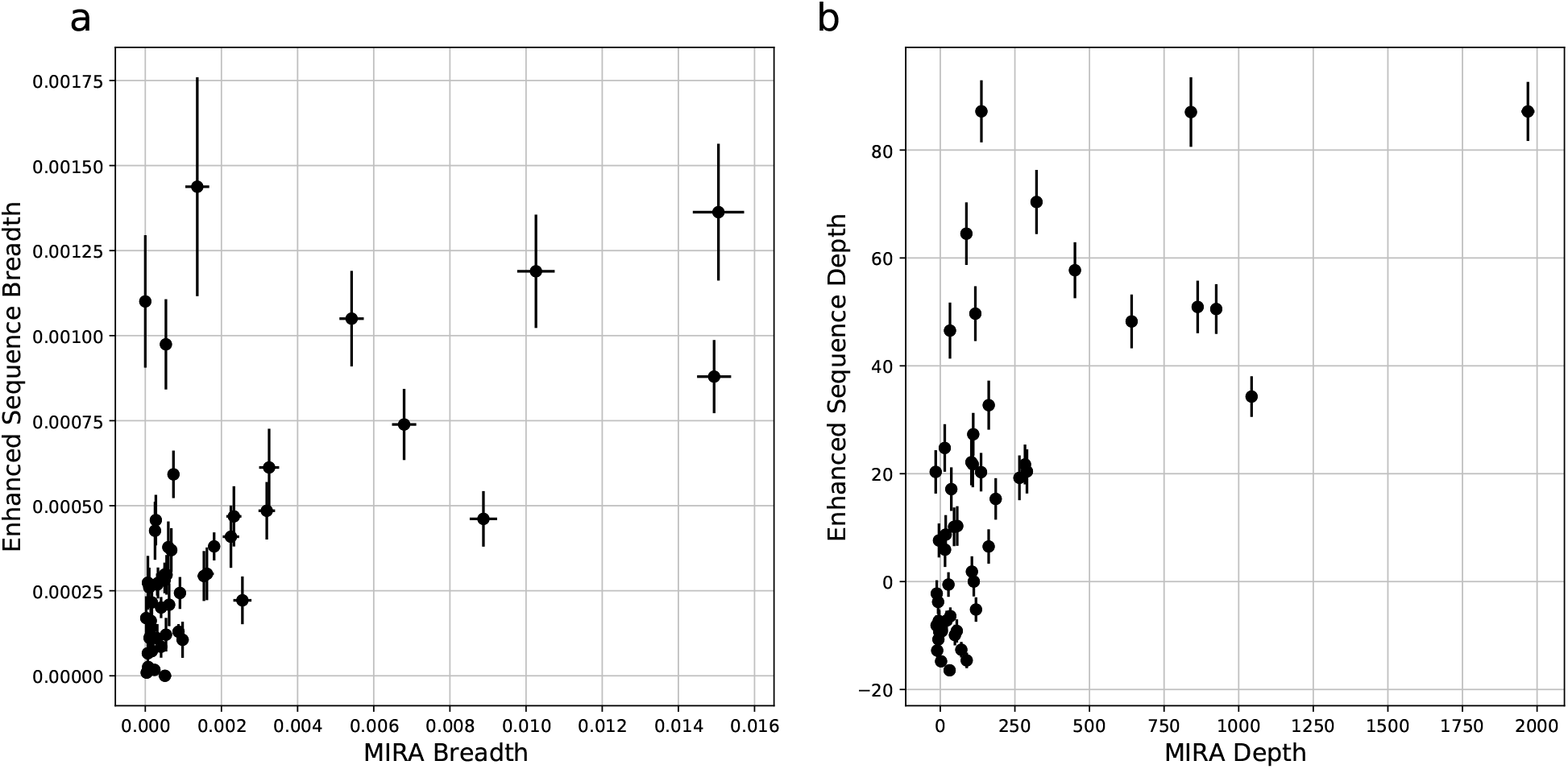
Clonal breadth and depth of the SARS-CoV-2 specific T-cell response can be estimated from MIRA-based profiling and from public enhanced sequences. Panel (a) is focused on breadth and shows a scatterplot of the relative fraction of the unique TCRβ DNA sequences in the repertoire that are assigned as MIRA or enhanced sequence TCRs. Panel (b) is focused on depth and shows a scatterplot of the summed logarithm of productive template counts across all SARS-CoV-2 associated clones from the two approaches, normalized by subtracting the logarithm of total template counts across all clones. In both panels, error bars on x and y represent the standard deviation.

MIRA-identified TCRs from an individual experiment are largely private (Supporting Figure S2), but the scale of data from MIRA should enable identification of antigen-specific TCR patterns that generalize to new individuals (Dash 2017; Glanville 2017). While those efforts advance, the high concordance between public enhanced sequences and MIRA defined breadth and depth provides a useful means of estimating these quantities in large populations.

### Analyzing T-cell response dynamics to SARS-CoV-2

As the T-cell response typically expands in the days following infection, then contracts to a steady memory state following clearance of viral antigens, the clonal breadth and depth should follow a similar trajectory. To test this hypothesis, the 1,015 COVID-19 subject samples were binned based on days since PCR-confirmed diagnosis with separate plots shown for the training and holdout sets used to discover this set of enhanced sequences (Figure 5). As expected, both breadth and depth indicate significant expansion of the T-cell response in the majority of subjects at time of diagnosis relative to healthy controls. As time progresses, both breadth and depth increase, reaching a peak in the 8-14 day and 15-28 day bins, then contracting slightly. Notably, both the 29-42 day and 43+ day bins show noticeably higher SARS-CoV-2-specific breadth and depth compared to controls, indicating the public enhanced sequences persist following presumed antigen clearance.

**Figure 5:**
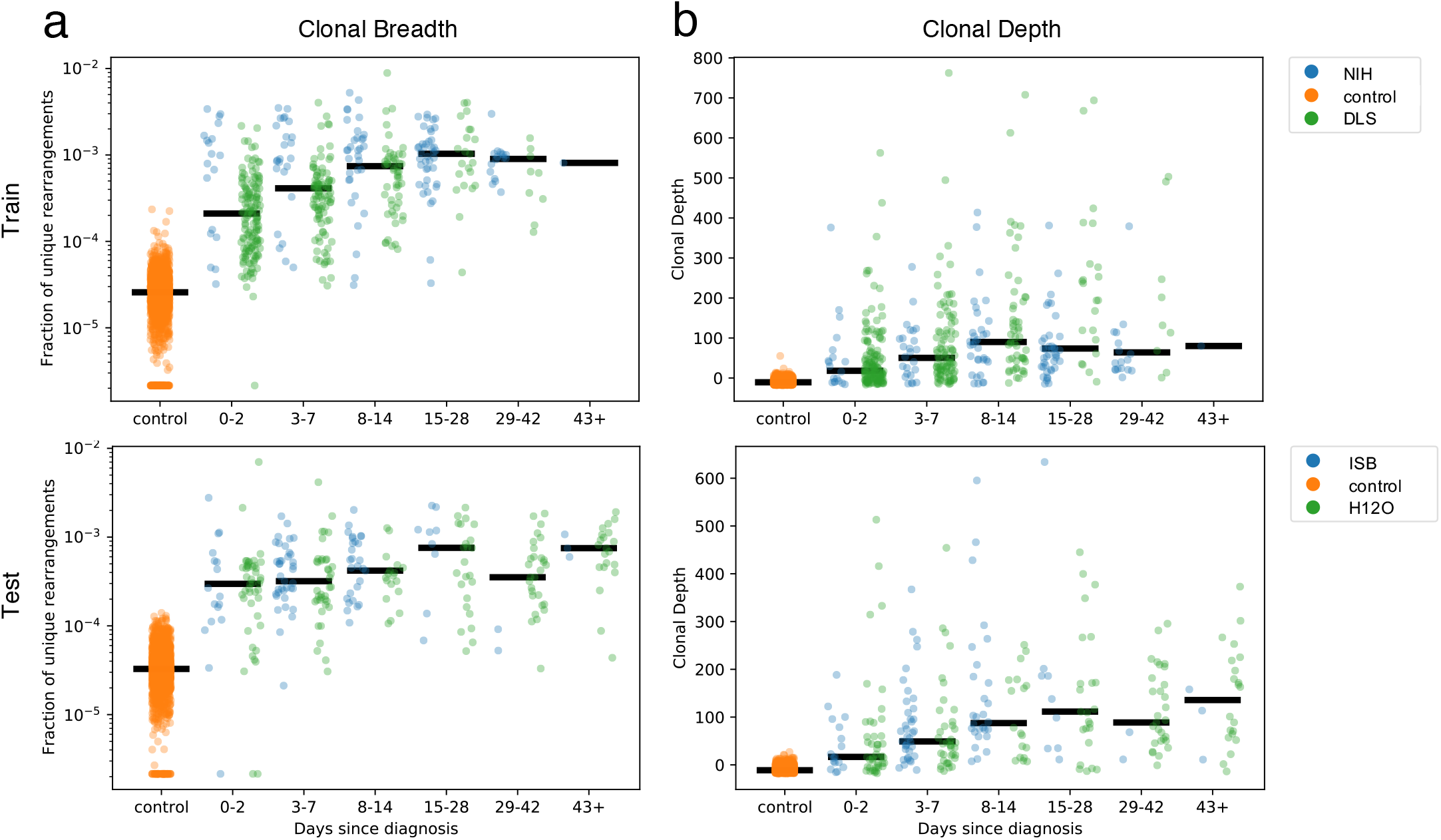
Breadth and depth of the immune response during SARS-CoV-2 infection and after recovery. Panels (a) and (b) represent, by time from diagnosis by a viral RT-PCR test, the clonal breadth (relative number of enhanced sequences observed) (a) and depth (a measure of frequency based on the summed logarithm of productive template counts normalized by subtracting the logarithm of total template counts) (b).

### Public enhanced TCR sequences are highly specific in diagnosing current and past SARS-CoV-2 infection

The significant expansion in SARS-CoV-2 specific clonal breadth and depth indicate that public enhanced sequences may constitute a useful biomarker for diagnosing past or present SARS-CoV-2 infection. Therefore, a simple logistic regression model was trained based on clonal breadth to separate cases from controls. As above, we initially used the DLS and NIH/NIAID cohorts, with a subset of controls, for model training and then tested on a holdout set of 325 samples from 276 distinct subjects from the ISB, H12O, and BWNW cohorts (with days from diagnosis information) and 1,702 pre-COVID-19 negative controls from other cohorts. Overall, the model was highly sensitive and specific in diagnosing current or past SARS-CoV-2 infection (Supporting Table S4). Using a target specificity of 99.8% across the 1,702 controls, the classifier demonstrates 77.4% sensitivity at 0-2 days post diagnosis (dpd) and 89.6% sensitivity at 3-7 dpd, further rising to 100% at 8-14 dpd. Notably, there is some reduced signal at 2-4 weeks from diagnosis; preliminary evidence suggests the negative cases are predominantly severe COVID-19 subjects who subsequently died or were in the ICU during the course of their illness, although further characterization with additional clinical / treatment data is required. The sensitivity for this first model is around 92-94% over a month after diagnosis (29+ dpd). We also investigated the model’s performance on later convalescent samples. From a separate set of 49 subjects whose blood was drawn ranging from 0-1 months, 1-2 months, and 2+ months from end of symptoms, there was ~90% sensitivity across all three of these time ranges suggesting a persistent T-cell signature after clearance of infection. The model performance is also robust to potential confounders such as age and sex (Supporting Figure S3).

Both the enhanced sequence identification and logistic regression parameter estimation should improve with additional training data. Therefore, using repertoires from an additional cohort, IRST, as well as additional data from the H12O and NIH/NIAID cohorts that are part of the version 003 ImmuneCODE release, we trained a new classifier with 1,421 unique cases and 2,447 controls. The cases include all the COVID-19 cohorts listed in Supporting Table S3 except for ImmuneRACE. This model used a variation of the prior training method that filters enhanced sequences present in difficult-to-classify controls in the training data, as described in the Methods. Five-fold cross validation was used to assess performance (Table 1). This model identifies a total of 4,242 enhanced sequences, more than double what was used in the initial model reported above, and increases sensitivity across multiple time ranges, reaching 85.1% at 3-7 days from initial diagnosis, 94.8% at 8-14 days from diagnosis, and >93% in all subsequent time bins analyzed out to 43+ days as shown in Table 1. For the set of 49 subjects whose blood was drawn ranging from 0-1 months, 1-2 months, and 2+ months from end of symptoms, sensitivity increased to 98%.

**Table 1:** Performance of a diagnostic model trained on all available data for 1,429 cases and 2,447 controls at 99.8% specificity. Performance reported for the subset of samples with annotated times since diagnosis or symptoms using five-fold cross validation.

| All @ 99.8% Specificity | % Sensitivity<br>(with 95% CI) | # Subjects |
| --- | --- | --- |
| Days since diagnosis: |  |  |
| 0-2 | 74.5 (69.3-79.1) | 329 |
| 3-7 | 85.1 (79.6-89.7) | 202 |
| 8-14 | 94.8 (90.8-98.4) | 134 |
| 15-28 | 93.6 (87.4-98.0) | 94 |
| 29-42 | 92.9 (86.4-98.5) | 70 |
| 43+ | 96.3 (90.2-100) | 54 |
| Days since end of symptoms: |  |  |
| 0-30 | 88.9 (66.7-100) | 9 |
| 31-60 | 100 (100-100) | 22 |
| 61+ | 100 (100-100) | 18 |

### Direct comparison of T-cell signature with antibody serology

To further explore the utility of this classifier, a direct comparison to serology was performed in the context of a real-world study. ImmuneRACE is a prospective virtual study that is enrolling individuals who were exposed to, actively infected with, or recovered from SARS-CoV-2 infection in at least 24 different geographic areas across the United States (Dines, 2020). After completion of an online consent and questionnaire, whole blood, serum, and a nasopharyngeal or oropharyngeal swab are collected by mobile phlebotomists.

From the first 100 subjects who reported SARS-CoV-2 infection by a viral RT-PCR test, immunoSEQ was performed from whole blood and serology assays were performed by LabCorp using two different tests: the multi-antibody test Elecsys^®^ Anti-SARS-CoV-2 (Roche) and the SARS-CoV-2 Antibody, IgG test (LabCorp). As shown in Table 2, 94 subjects (94%) were called positive by the T-cell classifier, while only 90 subjects (multi-serology) and 87 subjects (IgG only) were called positive by the serology assays. Treating the prior RT-PCR positive test as ground truth, these results suggest that the T-cell-based approach described here has greater positive percent agreement to RT-PCR results than antibody serology.

**Table 2:**
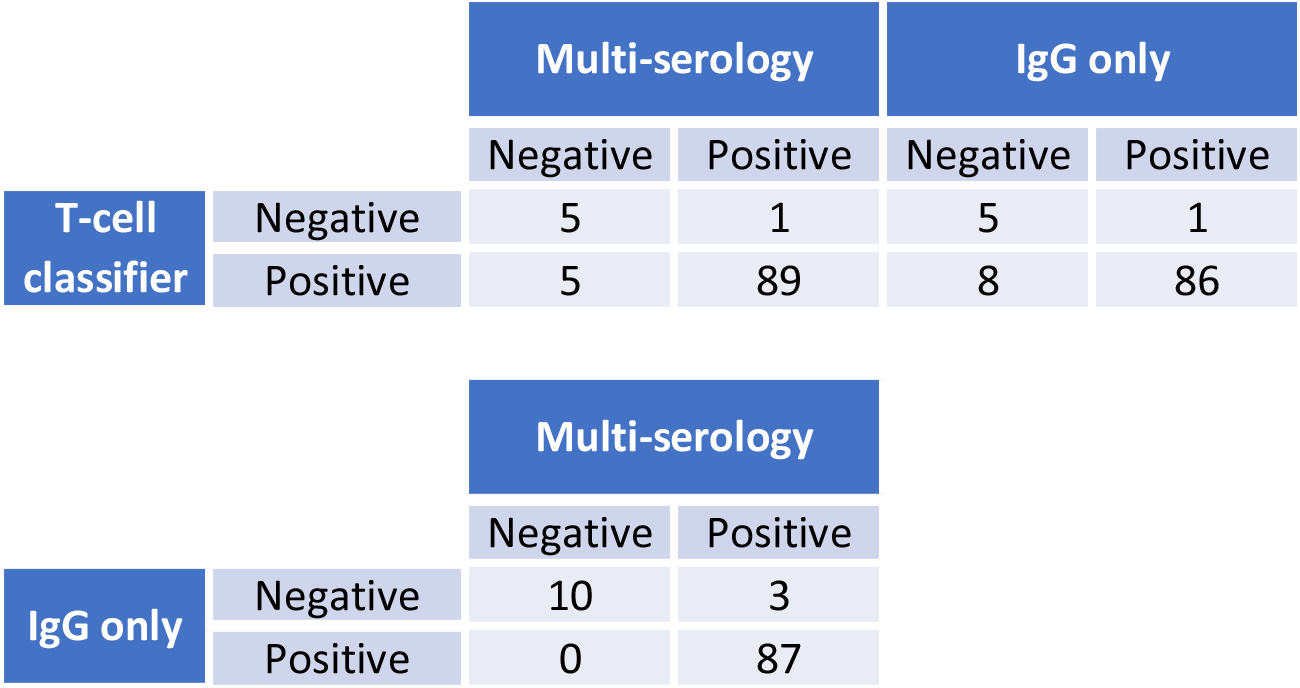
Relative performance of T-cell classifier versus antibody serology tests for 100 RT-PCR confirmed COVID-19 subjects. Three contingency tables show the agreement between each pair of tests, with the T-cell classifier having the most positive calls in agreement with RT-PCR results, followed by multi-antibody serology and then the IgG only test.

These 100 samples were collected between ~10 and 100 days from initial diagnosis, ranging from active infection through to convalescence. As there are reports of declining antibody signals over time, including reports of seroreversion (Sekine 2020, Peng 2020), we investigated whether the negative serology or T-cell diagnostic calls have any specific time-based trends. No significant associations with days from diagnosis (Supporting Figure S4) were seen for either approach. In evaluating differences in the testing results, we also considered potential differences in disease severity. Nearly all subjects in this comparison had multiple symptoms from COVID-19, but there was one asymptomatic subject who had two positive PCR tests days apart. This subject tested negative by both antibody serology tests but was positive by the T-cell-based classifier.

We also characterized the first 23 subjects from the “exposed” cohort from ImmuneRACE, who at the time of study enrollment reported exposure to someone with SARS-CoV-2 infection but themselves were not actively symptomatic nor diagnosed with COVID-19. As a result of the virtual study design, sample collection occurred several days to weeks following enrollment, allowing for several individuals to progress to acute infection prior to sample collection. A comparison between immunoSEQ and two antibody serology tests (same as above) was performed on these 23 subjects to determine whether antibody (B cell) or T-cell signals were present in these subjects. For serology, one subject was positive by the IgG-only assay, but none by the multi-isotype assay. In comparison, two different subjects were called positive based on the T-cell classifier, but negative by both antibody serology assays tested. Medical record review of these two subjects confirmed that they both had developed COVID-19 and had a positive RT-PCR test 4 days prior to the time of sample collection, with one subject reporting symptoms 5 days prior. While limited in total number, these results suggest that the T-cell classifier may be more capable of detecting signs of SARS-CoV-2 infection earlier, and in less severe cases, than tests that detect antibody (B cell) response.

## Discussion

We have described an approach that uses fine mapping of TCR sequences to hundreds of antigens in conjunction with statistical association of over a thousand public enhanced sequences to track the breadth and depth of the cellular immune response to SARS-CoV-2. This immunoSEQ T-MAP COVID approach utilizes a small volume (1-2 milliliters) of whole blood and is compatible with most standard collection methods. It reliably and reproducibly identifies and tracks SARS-CoV-2 specific T-cell clones soon after infection and for months after recovery for most subjects based on currently available samples and data. The map for CD4 T cells is currently being generated through the same combination of MIRA and case/control experiments and will be reported on alongside our ImmuneCODE public data resource.

There are many advantages of the molecular assay presented as compared to standard techniques such as ELISpot for assessing cellular immune response. The biggest advantage is that standard functional assays require live cells and the results vary depending on how the sample was handled, stored and transported. The T-cell molecular assay used here is based on DNA, which is highly stable, and probes T cells with resolution down to 1/1,000,000 cells whereas functional assays are usually only sensitive down to 1/10,000 cells. The approach assesses T cells sampled randomly from blood and, unlike functional assays, is not restricted to reagent-limited subcompartments of the cellular immune response.

Although functional T-cell assays are challenging to perform, in the hands of experts their use has led to many important findings about the cellular immune response to SARS-CoV-2. This includes early profiles of the immunoreactivity of different pools of SARS-CoV-2 antigens to CD4 and CD8 T cells and identification of potential cross-reactive T cells to SARS-CoV-2 in healthy individuals (Grifoni 2020, Weiskopf 2020, Nelde 2020, Ferretti 2020). Other studies have revealed strong associations between the T-cell response and disease severity (for review, see Vabrat 2020 and Chen 2020). Evidence has also emerged in a number of independent studies demonstrating detectable T-cell responses in PCR-confirmed individuals in mild or asymptomatic cases where serology was not initially detected or in those who later serorevert (Sekine 2020, Peng 2020).

This manuscript recapitulates some of these findings while also adding greater scale and resolution to the emerging picture of the T-cell response. While other antigen-stimulation approaches provide an aggregated result for how a pool of antigens may respond, MIRA allows for the simultaneous characterization of hundreds to thousands of individual antigen addresses, associating tens of thousands of TCRs to specific antigens. Here we also demonstrated that through population scale sequencing of immune repertoires, public TCR sequences to SARS-CoV-2 can be identified that collectively shed light on the shared immune response at the population level. These sequences, in combination with the MIRA data, allow characterization of disease-and antigen-specific responses including the breadth and depth of the overall cellular response to a viral infection.

Assessing T-cell responses still comes with challenges that will be addressed in future work. As previously discussed, the MIRA results described here assess the CD8 T-cell response and further work (underway) is needed to characterize the CD4 T-cell response. Also, despite including several hundred peptides in our initial CD8 T-cell panels, including some of the strongest predicted binders, these likely represent a fraction of the antigens presented in different HLA contexts. HLA diversity is a key part of the adaptive immune response; we have used large, diverse study cohorts to account for this variation, but continue to collect additional data in an effort to fully characterize rare HLA alleles.

The importance of characterizing the cellular immune response has applications in development of therapeutics and vaccines as well as in clinical evaluation of exposure or response to SARS-CoV-2. One potential translational application of this approach is to identify and track the T-cell response against immunogenic, virus-specific epitopes as a possible correlate of protection. Our results suggest that natural immune responses to SARS-CoV-2 include responses to current targets of vaccines in development, such as the surface glycoprotein (spike), but also include strong or in some cases stronger responses to antigens from other viral proteins depending on HLA context, consistent with other reports (Grifoni 2020, Ferretti 2020). Another application for this approach is as a T-cell-based diagnostic to identify individuals with recent or past infection. The data presented here suggest frequent and persistent TCRs are elevated as far out as ~100 days, which is currently the furthest timepoint we have assessed. This supports the development of a TCR-based clinical diagnostic to broadly identify past exposure, especially in individuals in whom an antibody response is delayed, is muted, or wanes. The direct comparison of the T-cell response to antibody serology described here, together with a growing body of evidence indicating T cells persist beyond antibody responses, support the clinical utility of a Tcell-based approach to detect immune responses to SARS-CoV-2.

The scientific community has rapidly developed and deployed many tools to characterize the immune response to SARS-CoV-2 in an effort to aid the development of diagnostics and treatments for COVID-19. Future success in controlling and containing the current pandemic will rely on a complete picture of the biology of disease and treatment response. The development of a reproducible, high-throughput, high-resolution molecular approach to assess the T-cell response will serve to fill an important unmet need in characterizing the adaptive immune response to exposure to SARS-CoV-2 antigens.

## Data Availability

The COVID-19 MIRA data and COVID-19 study immunosequencing data are freely available for analysis and download from the Adaptive Biotechnologies immuneACCESS site at https://clients.adaptivebiotech.com/pub/covid-2020

## Data Availability

As part of the ImmuneCODE data resource (Nolan 2020), the COVID-19 MIRA data and COVID-19 study immunosequencing data are freely available for analysis and download from the Adaptive Biotechnologies immuneACCESS site under the immuneACCESS Terms of Use at https://clients.adaptivebiotech.com/pub/covid-2020.

## Acknowledgements

The ImmuneCODE data resource that underlies this analysis paper is the result of collaboration between many individuals and organizations working together to advance global understanding of SARS-CoV-2 and COVID-19. We are grateful for the support and participation of all our partners. We are especially grateful for the generosity of the participants who donated blood for this and other studies.

These study data would not be available if not for the hard work of the entire Adaptive Biotechnologies laboratory and support staff who rose to solve the new challenges posed by the pandemic; we cannot thank this incredible team enough.

From Bloodworks Northwest (Seattle, WA), we would like to thank Caitlin Jirovsky, Matthew Bird and Rohit Nariya for operational involvement and Evan Delay, Adam Skrzekut and Dr. David Lin for oversight and management.

For the ImmuneRACE study, we would like to thank Covance/LabCorp and Illumina for their ongoing partnership.

We would also like to thank Ted Meeds, Elon Portugaly, Bin Shao, Leo Xia, and many others for helpful discussions.

## Methods

### Clinical sample collection

Samples were collected based on each institution’s study protocol, as reviewed by their Institutional Review Board. From all sources, whole blood samples were collected in K2EDTA tubes and were stored until being shipped to Adaptive as frozen whole blood, isolated PBMC or DNA extracted from either blood or PBMC for immune profiling analyses via the immunoSEQ Assay and/or MIRA.

Samples provided by the NIAID were collected under approval by Comitato Etico Provinciale (protocol NP-4000), and by Comitato Etico, Ospedale San Gerardo Monza (protocol COVID-STORM). The Brescia study includes collection of discarded blood samples obtained from patients who were admitted at ASST Spedali Civili Brescia following positive nasopharyngeal swab for SARS-CoV-2 infection. Samples were obtained from all patients admitted to the hospital, as long as discarded material was available. Patients in Monza were enrolled when they were admitted to the San Gerardo Hospital in Monza, criteria for enrollment required a positive COVID-19 PCR test.

Samples provided by Hospital 12 de Octubre were collected under approval by Comite Etico del Hospital 12 de Octubre, Madrid IC (protocol 20/161). Participants were recruited at Hospital Universitario 12 de Octubre from inpatient and hospital workers with a positive COVID-19 PCR test.

Samples provided by Swedish-ISB were collected under approval by the Providence St. Joseph’s Health system IRB (STUDY2020000175). Study participants were recruited at clinics associated with Swedish Medical Center with a confirmed diagnosis by SARS-CoV-2 PCR or persons under investigation (with PCR pending) with >3 diagnostic criteria. SARS-CoV-2 PCR was performed at enrollment to confirm diagnosis.

Samples provided by IRST and AUSL Romagna were collected under approval by CEROM (IRSTB113). Specifically, remnants of whole blood samples from diagnostic procedures of SARS-CoV-2 nasopharyngeal swab positive patients were stored frozen at -20°C before shipment to Adaptive.

Whole blood samples from DLS (Discovery Life Sciences, Huntsville, AL) were collected under Protocol DLS13 for collection of remnant clinical samples. All DLS subjects had tested positive for SARS-CoV-2 viral exposure by an Abbott RealTime SARS-CoV-2 RT-PCR assay.

From Bloodworks Northwest (Seattle, WA), volunteer donors recovered from COVID-19 were consented and collected under the Bloodworks Research Donor Collection Protocol BT001. Samples were processed for PBMC and donor data reported by the Biological Products division of Bloodworks NW under standard operating procedures. Inclusion criteria for samples collected by Bloodworks included age of at least 18 years old, weight of more than 110 lbs, a diagnosis of SARS-CoV-2 infection, at least 28 days since positive screening or days since last symptoms or a negative SARS-CoV-2 PCR test, and a provision of informed consent to participate in the study.

Controls were selected from primarily healthy controls drawn before 2020 by Diagnostic Laboratory Services, as well as other non-COVID studies. These samples are presumed negative and include collections during seasons with high prevalence of vaccination against, and/or infection with, the influenza A/B viruses and seasonal coronavirus(es) in order to exclude potential cross-reactivity.

### ImmuneRACE sample collection and serology testing

The ImmuneRACE study (Dines, 2020) is a prospective, single group, multi-cohort, exploratory study of participants exposed to, infected with, or recovering from COVID-19 (NCT04494893). Participants from across the United States were consented and enrolled via a virtual study design, with cohorting based on participant-reported clinical history following the completion of both a screening survey and study questionnaire. All participants provided informed consent for sample collection and metadata use. Whole blood, serum, and a nasopharyngeal or oropharyngeal swab were collected from participants by trained mobile phlebotomists. The study was approved by Western Institutional Review Board (WIRB reference number 1-1281891-1, Protocol ADAP-006).

In this research, samples were selected from the first 100 individuals with self-reported COVID-19 based on an RT-PCR SARS-CoV-2 test from the acute and recovered cohorts as well as 23 individuals from the exposed cohort who at the time of enrollment and study questionnaire were within 2 weeks of exposure to someone diagnosed with COVID-19, asymptomatic, and not diagnosed with COVID-19. Whole blood samples were processed identically to other studies for the immunoSEQ assay. Additionally, serum samples were tested by Covance/LabCorp using two different EUA approved assays: 1) Elecsys® AntiSARS-CoV-2; Roche: qualitative detection of high affinity antibodies to SARS-CoV-2 including all isotypes, but preferentially detects IgG antibodies (https://www.labcorp.com/tests/164068/sars-cov-2antibodies); and 2) SARS-CoV-2 Antibody, IgG; LabCorp: qualitative detection of IgG antibodies to SARSCoV-2 (https://www.labcorp.com/tests/164055/sars-cov-2-antibody-igg).

### Viral peptide selection

Using the NCBI genome reference for SARS-CoV-2 (RefSeq accession: NC_045512.2), a list of candidate 9-10AA long peptides from across the whole viral genome was identified based on predicted affinity (<1% rank) using NetMHCpan version 4.1 (Andreatta 2016; Nielsen 2003) to common HLA-A and -B alleles as determined in the Allele Frequency Net Database (Gonzalez-Galarza 2020). An additional 121 peptides were added to this list from (Ahmed 2020), which identified candidate epitopes conserved between SARS-CoV-1 and SARS-CoV-2 and optimized for global HLA coverage. The final set of peptides included candidate epitopes for most common HLA alleles across the globe: A*01:01, A*02:01, A*02:07, A*03:01, A*11:01, A*23:01, A*24:02, A*31:01, A*33:01, A*33:03, A*68:01, B*07:02, B*08:01, B*13:01, B*15:01, B*15:02, B*18:01, B*27:05, B*35:01, B*40:01, B*44:02, B*46:01, B*51:01, B*58:01, C*14:02, C*15:02. Peptides were synthesized by GenScript (Piscataway, NJ). The complete list of peptides is in Table S1.

The 545 peptides were then pooled in a combinatorial fashion as described previously (Klinger 2015); peptides that were overlapping or in close proximity in the viral proteome were grouped together into antigen sets. Each antigen set was then placed in a subset of 6 unique pools out of 11 pools; hereto after referred to as its occupancy. In order to estimate an empirical false discovery rate and gauge assay quality, we purposefully left > 40% of the unique occupancies empty to assess the rate at which clones are spuriously sorted and detected in 6 pools with no query antigen present.

Phylogenetic context of candidate epitopes was assessed using a customized BLAST database of 55 RefSeq coronavirus genomes across the Coronaviridae family (Sayers 2019). BLAST searches were optimized for short sequence queries using the “-task blastp-short” argument and all full-length, exact matching TCRs were used to assess the phylogenetic placement of each candidate epitope. Using the taxonomic annotations available from the NCBI taxonomy browser, the most recent common ancestor was defined as the most recent taxonomic node shared by all terminal taxa that shared an exact match to the epitope. Each epitope was also assessed for its homology to each of 4 endemic human coronaviruses: Human coronavirus 229E, Human coronavirus HKU1, Human coronavirus NL63, and Human coronavirus OC43 in order to explore the role of cross reactivity in T cell responses.

### Antigen stimulation experiments (MIRA)

Antigen-specific TCRs were identified using the Multiplex Identification of T-cell Receptor Antigen Specificity (MIRA; Klinger 2015). For these MIRA from 61 COVID-19 subjects, T cells from PBMC were first expanded with anti-CD3 (Biolegend clone OKT3, San Diego, CA) at 30 ng/ml, IL-2 (Biolegend, San Diego, CA) at 20 ng/ml, and IL-15 (Biolegend, San Diego, CA) at 5 ng/ml for 8-13 days. Expanded memory cells were then stimulated by peptide pools at 37°C for ~18 hours. Replicate wells of cells were harvested from the culture and pooled and then stained with antibodies for analysis and sorting by flow cytometry. Cells were then washed and suspended in PBS containing FBS (2%), 1mM EDTA and 4,6-diamidino-2-phenylindole (DAPI) for exclusion of non-viable cells. Cells were acquired and sorted using a FACS Aria (BD Biosciences) instrument. Sorted antigen-specific (CD3+CD8+CD137+) T cells were pelleted and lysed in RLT Plus buffer for nucleic acid isolation. Analysis of flow cytometry data files was performed using FlowJo (Ashland, OR).

RNA was then isolated using AllPrep DNA/RNA mini and/or micro kits, according to manufacturer’s instructions (Qiagen). RNA was reverse transcribed to cDNA using Vilo kits (Life Technologies), and TCRβ amplification performed using the immunoSEQ assay described below.

After immunosequencing, we examined the behavior of T-cell clonotypes by tracking read counts across each sorted pool. True antigen-specific clones should be specifically enriched in a unique occupancy pattern that corresponds to the presence of one of the query antigens in 6 pools. We have reported on methods to assign antigen specificity to TCR clonotypes previously (Klinger 2015). In addition to the previously published methods, we also developed a non-parametric Bayesian model to compute the posterior probability that a given clonotype is antigen specific. This model uses the available read counts of TCRs to estimate a mean-variance relationship within a given experiment as well as the probability that a clone will have zero read counts due to incomplete sampling of low frequency clones. Together, this model takes the observed read counts of a clonotype across all 11 pools and estimates the posterior probability of a clone responding to all possible 11 choose 6 addresses and an additional hypothesis that a clone is activated in all pools (truly activated, but not specific to any of our query antigens). To define antigen specific clones, we identified TCR clonotypes assigned to a query antigen from this model with a posterior probability ≥ 0.7.

### Immunosequencing of TCR repertoires

For blood or PBMC samples, genomic DNA was extracted from either peripheral blood mononuclear cells or from peripheral blood samples using the Qiagen DNeasy Blood Extraction Kit (Qiagen). As much as 18 μg of input DNA was then used to perform immunosequencing of the CDR3 regions of TCRβ chains using the ImmunoSEQ Assay. Briefly, input DNA was amplified in a bias-controlled multiplex PCR, followed by high-throughput sequencing. Sequences were collapsed and filtered in order to identify and quantitate the absolute abundance of each unique TCRβ CDR3 region for further analysis as previously described (Robins 2009, Robins 2012, Carlson 2013). In order to quantify the proportion of T cells out of total nucleated cells input for sequencing, or T cell fraction, a panel of reference genes present in all nucleated cells was amplified simultaneously (Pruessmann 2020).

### Characterization of the T-cell response with MIRA

In two separate analyses, each subject’s response to the antigens presented by the MIRA panel was summarized by the fraction of T cells responding to each protein, or to each antigen. Donors were clustered with average-linkage hierarchical clustering into five clusters (number of clusters chosen by visual inspection). For antigen-based clustering, only the 50 antigens present in the largest numbers of donors were used. 47 of the 61 donors, spread across the three large clusters, had HLA typing available. Association of each HLA with each antigen-based cluster was assessed with a one-sided Fisher’s Exact Test, using all available HLA typing.

### Enhanced TCR Sequence Discovery and Classification from Case / Control studies

Public TCRβ amino acid sequences (“enhanced sequences”) were associated with SARS-CoV-2 infection as described previously (Emerson 2017). Briefly, one-tailed Fisher’s exact tests were performed on all unique TCR sequences comparing the presence in SARS-CoV-2 positive samples with negative controls. Unique sequences were defined by their V gene, J gene, and CDR3 amino acid sequence. For subjects with longitudinal sampling, only the latest available sample was used.

Enhanced sequences were turned into a classifier predicting current or past infection with SARS-CoV-2 using a simple two feature logistic regression with dependent variables E and N, where E is the number of unique TCRβ DNA sequences that encode an enhanced sequence and N is the total number of unique TCRβ DNA sequences in that subject.

The significance threshold used to define the enhanced sequence set was chosen to maximize out-of-sample classification accuracy using 5-fold cross validation. In all cases described, the model identified p<0.001 as an optimal threshold, though the results were largely insensitive to the specific threshold chosen (data not shown).

In the final diagnostic classifier, an additional step was added to filter enhanced sequences that were common in the negative control samples as follows: first, a model was built with the initial set of enhanced sequences. Predictions were made on the training set to identify false positive control samples (model score > .35). Sequences that were present in two or more false positive control samples were removed from the enhanced sequence set before the final model was trained. The number of control samples a sequence was present in before exclusion and the score threshold for defining false positives were obtained by maximizing out-of-sample classification accuracy using 5-fold cross validation.

### The breadth and depth of a disease-specific T-cell response

To summarize the extent to which a set of sequenced T cells is specific to a disease or set of antigens, we define the quantities clonal breadth *B* and clonal depth *D* as follows. For a given repertoire *j*, let *N_j_* be the number of unique TCR DNA sequences in the repertoire; *t_ij_, i =* 1*,…,N_j_*, be the estimated number of T cells that have TCRβ DNA sequence *i* (assumed to derive from the same progenitor cell); and *M_j_ =∑_i_ t_ij_* be the total number of T cells sequenced by the assay.

Then, for a given set of sequences 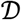, the *clonal breadth of j with respect* to 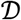 is defined to be 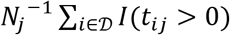, where *I* is the indicator function and the summation is over all clones in 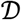. That is, clonal breadth is the proportion of lineages in the repertoire that are mapped to the disease as defined by 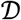.

Clonal depth is similar, but attempts to capture the extent of clonal expansion of each lineage. Because the observed number of DNA templates derived from the same progenitor clone, *t_ij_*, is the result an exponential growth process, we use as our base measure of depth a number that is proportional to the estimated number of clonal generations that lineage *i* went through, *g_ij_ =* log*_2_*(1 *+ t_ij_*). Then the *clonal depth of j with respect to* 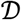 is defined to be 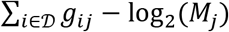, which estimates the relative number of clonal expansion generations across the TCRs in 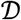, normalized by the total number of TCRs sequenced in the assay.

Error estimates on clonal breadth are derived starting from the assumption of Poisson error on the counting statistics comprising both the numerator 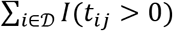 and denominator *N_j_*. For clonal breadth, the full error on the quotient quantity 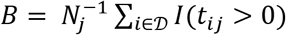 is then given by 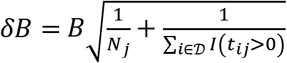.

For clonal depth, errors are estimated starting from the same assumption of Poisson counting errors on both template counts for individual clones *t_i_* as 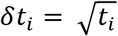 and on total templates *M_j_* as 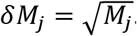. This error is then propagated along to *g_ij_* as 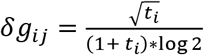. Adding in quadrature the errors on the *g_ij_* along with the error on the normalization term 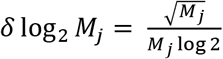 gives the final uncertainty in the depth as 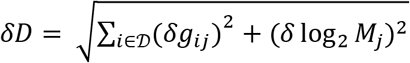.

**Supporting Figure S1:**
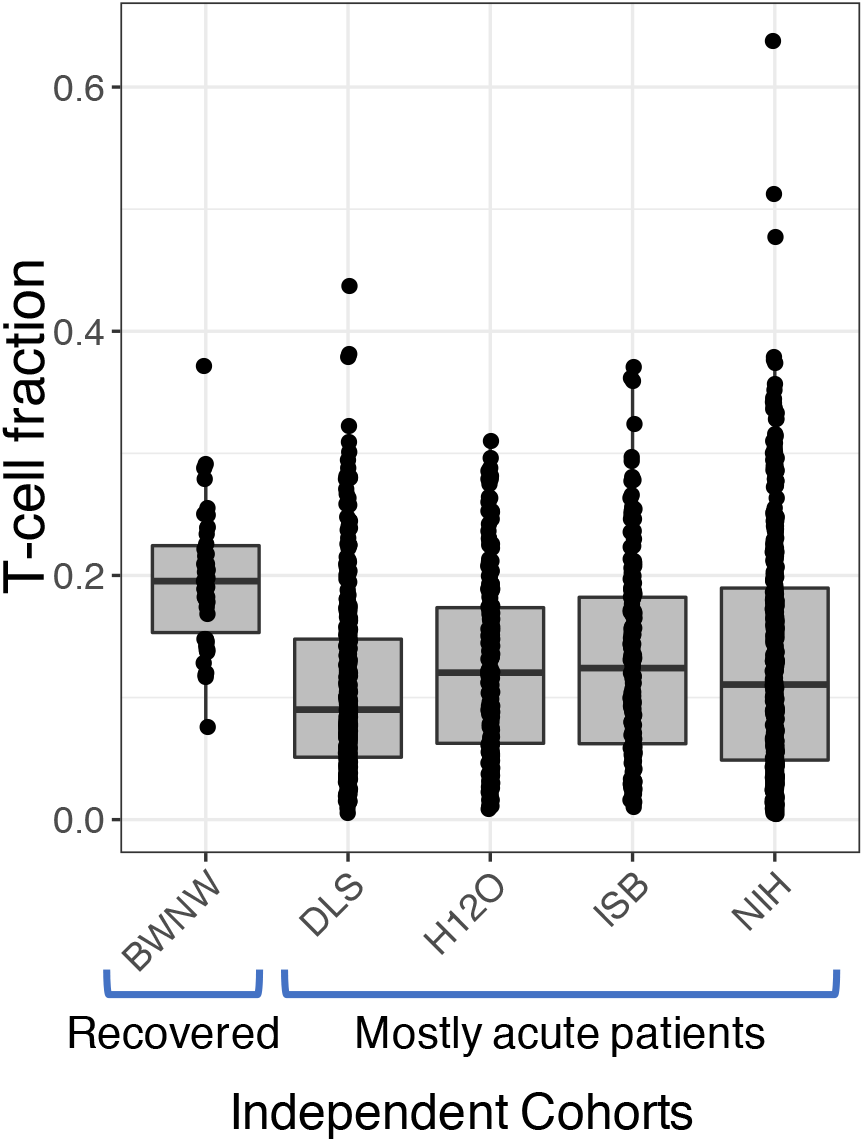
Distribution of T-cell fraction across different COVID-19 cohorts. While the samples from BWNW (exclusively convalescent subjects) are near normal values, the overall T-cell fraction is depressed across all other cohorts, which are enriched for acutely infected subjects. Individuals on the low end of these distributions would be considered severely lymphopenic.

**Supporting Figure S2:**
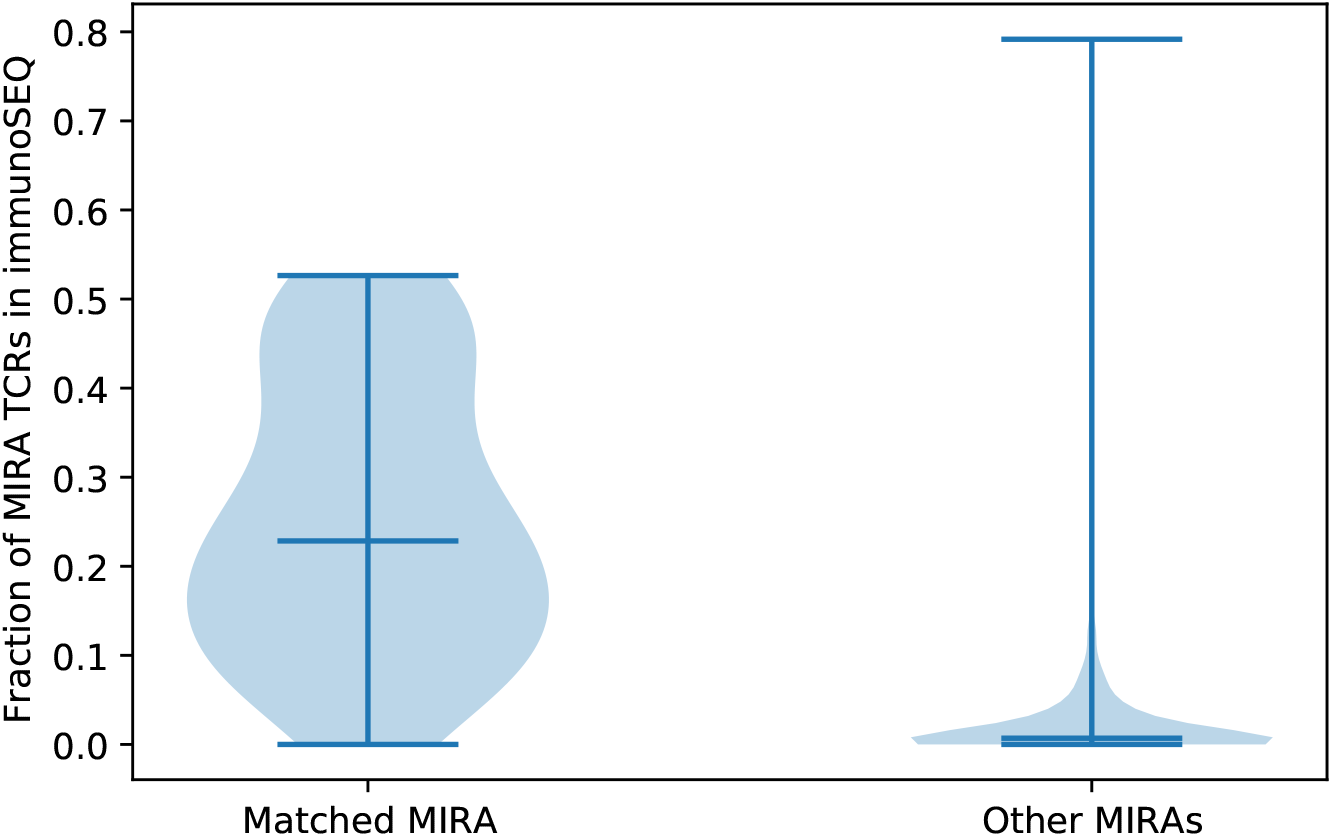
Overlap of MIRA with immunoSEQ (within and across subject). Within individuals, a median of about 25% of the TCRs identified by MIRA are detectable in a separate sample assessing the overall immune repertoire. Across individuals, this comparison drops much lower suggesting that a majority of the detectable response is due to private TCRs.

**Supporting Figure S3:**
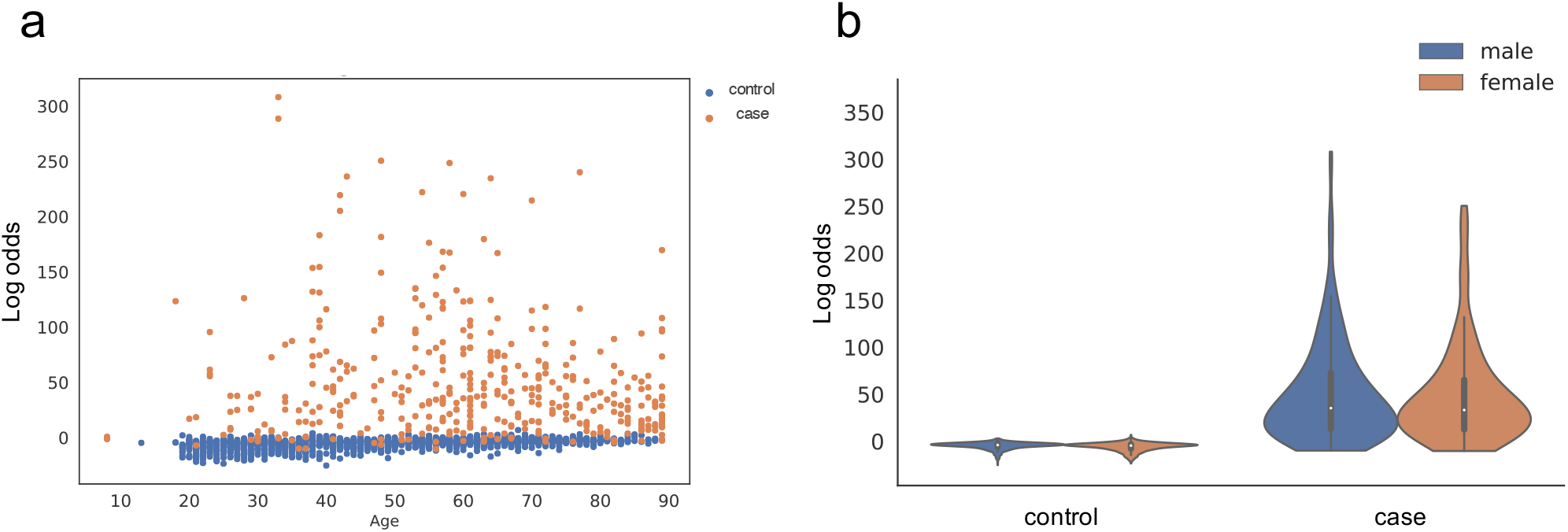
Model predictions separate SARS-CoV-2 cases from controls across ages (a) and in both males and females (b). Both plots report model scores as the untransformed log-odds estimated from the logistic regression classifier. The violin plot in panel (b) visualizes the density of log-odds scores among male and female cases and controls, with median and interquartile range values indicated.

**Supporting Figure S4:**
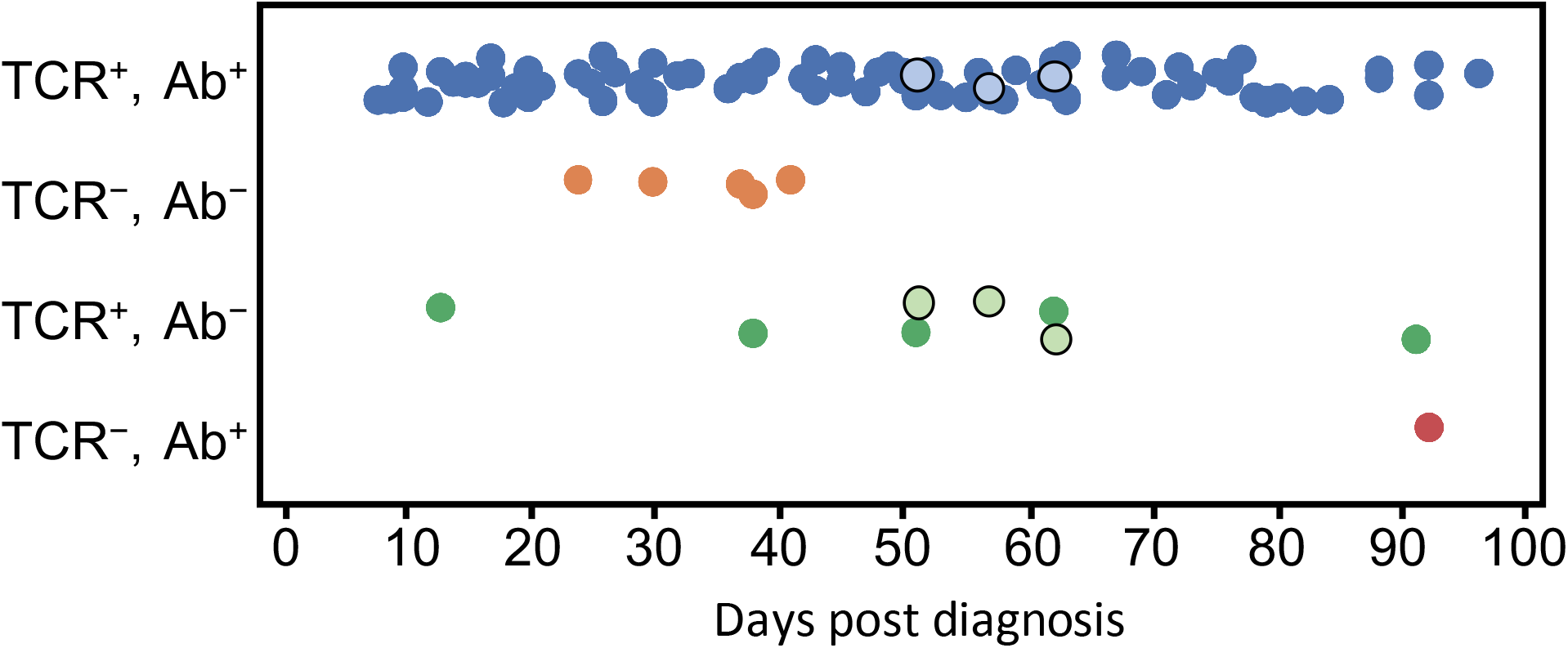
Performance by time since diagnosis for the T-cell classifier and antibody serology tests for 100 RT-PCR confirmed COVID-19 subjects. The three outlined points represent samples where the multi-antibody serology test was positive but IgG only was negative, changing the category of the points depending on which antibody test is being compared. No significant associations with time are observed for the negative calls from either the T-cell classifier or the antibody tests.

*Supporting tables S1 and S2 are available as an Excel file on the publisher’s website*.

Supporting Table S1: Complete list of antigen locations and peptides with matches between the MIRA experiments, as well as any exact sequence matches to enhanced sequences identified in the initial case/control study.

Supporting Table S2: List of antigens from MIRA data where putative HLA restrictions can be attributed based on using a Mann-Whitney’s U test over the number of mapped TCRs per experiment.

**Supporting Table S3:** Summary of Clinical Cohorts included in this study, including summaries of demographic parameters.

| Name of cohort | Sample Count | Subject Count | Institution | Age | % Male | Study description |
| --- | --- | --- | --- | --- | --- | --- |
| COVID-19-BWNW | 62 | 62 | Bloodworks Northwest | 54 (20, 79) | 52 | Whole blood samples from convalescent subjects collected at Bloodworks Northwest (Seattle, WA) |
| COVID-19-DLS | 431 | 337 | Discovery Life Sciences | 70 (23, 89) | 49 | Whole blood samples collected during routine patient care in acute and convalescent phases procured through Discovery Life Sciences (Huntsville, AL) |
| COVID-19-ISB | 157 | 114 | Institute for Systems Biology | 61 (18, 89) | 42 | Whole blood samples collected under the INCOVE project at Providence St. Joseph Health (Seattle, WA). Subjects were enrolled during the active phase and monitored through disease |
| COVID-19-NIH/NIAID | 389 | 285 | National Institute for Allergy and Infectious Diseases (NIAID) | 45 (29, 89) | 58 | Whole blood samples were collected in Brescia and Monza (Italy) during active infection, and provided to the NIAID (Bethesda, MD) for DNA extraction |
| COVID-19-H12O | 612 | 570 | Hospital Universitario 12 de Octubre | 58 (8, 89) | 29 | Whole blood samples were collected at the Hospital Universitario 12 de Octubre (Madrid, Spain) during the active or convalescent phase |
| COVID-19-IRST | 64 | 53 | Istituto Scientifico Romagnolo per lo Studio e la Cura dei Tumori (IRST) / AUSL-Romagna | 78 (20, 89) | 49 | Whole blood samples were collected by IRST/AUSL (Romagna, Italy) during active infection |
| COVID-19-ImmuneRACE | 123 | 123 | Adaptive Biotechnologies | 43 (18, 74) | 27 | Whole blood samples were collected from subjects from 24 geographic areas in the US with active infection, in convalescent phase, or exposed to SARS-CoV-2 |

**Supporting Table S4:** Performance of a diagnostic model trained on an initial data set from two independent sources and tested on a hold-out data set of 276 distinct case samples and 1,702 pre-COVID-19 controls. Performance is reported at a level of 99.8% specificity for the classifier.

|  | Holdout |  | Train (5x cross validation) |  |
| --- | --- | --- | --- | --- |
|  | % Sensitivity<br>(with 95% CI) | # Subjects | % Sensitivity<br>(with 95% CI) | # Subjects |
| Days since diagnosis: |  |  |  |  |
| 0-2 | 77.4 (65.7-87.8) | 63 | 57.3 (49.7-65.5) | 164 |
| 3-7 | 89.6 (81.2-95.4) | 79 | 78.3 (71.3-86.1) | 125 |
| 8-14 | 100 (100-100) | 46 | 88.4 (81.2-94.5) | 88 |
| 15-28 | 81.8 (68.4-95.2) | 33 | 90.5 (83.8-98.3) | 64 |
| 29-42 | 93.5 (83.3-100) | 31 | 100 (100-100) | 26 |
| 43+ | 91.7 (78.6-100) | 24 | 100 (NA-NA) | 1 |
| Days since end of symptoms: |  |  |  |  |
| 0-30 | 88.9 (62.5-100) | 9 |  |  |
| 31-60 | 90.9 (76.9-100) | 22 |  |  |
| 61+ | 100 (80.9-100) | 18 |  |  |

## References

1. Ahmed SF, Quadeer AA, McKay MR. 2020. Viruses 12(3):254. Preliminary Identification of Potential Vaccine Targets for the COVID-19 Coronavirus (SARS-CoV-2) Based on SARS-CoV Immunological Studies.

2. Altmann DM and Boyton RJ. 2020. Sci. Immunol. 10.1126/sciimmunol.abd6160. SARS-CoV-2 T cell immunity: Specificity, function, durability, and role in protection.

3. Andreatta M, Nielsen M. 2016. Bioinformatics 32(4);511–7. Gapped sequence alignment using artificial neural networks: application to the MHC class I system.

4. Cao X. 2020. Nature Reviews Immunology 20:269-270. COVID-19: immunopathology and its implications for therapy.

5. Carlson CS, Emerson RO, Sherwood AM, Desmarais C, Chung M, Parsons JM, Steen MS, LaMadrid-Herrmannsfeldt MA, Williamson D, Livingston RJ, Wu E, Wood BL, Rieder MJ, Robins HS. 2013. Nature Communications 4:2680. Using synthetic templates to design an unbiased multiplex PCR assay.

6. Chen Z, Wherry EJ. 2020. Nature Reviews Immunology. https://doi.org/10.1038/s41577-0200402-6. T cell responses in patients with COVID-19.

7. Dash P, Fiore-Gartland AJ, Hertz T, Wang GC, Sharma S, Souquette A, Crawford JC, Clemens EB, Nguyen THO, Kedzierska K, La Gruta NL, Bradley P, Thomas PG. 2017. Nature. Jul 6;547(7661):89-93. Quantifiable predictive features define epitope-specific T cell receptor repertoires.

8. DeWitt WS III, Emerson RO, Lindau P, Vignali M, Snyder TM, Desmarais C, Sanders C, Utsugi H, Warren EH, McElrath J, Makar KW, Wald A, Robins HS. 2015. J Virol. 89(8):4517-26. Dynamics of the cytotoxic T cell response to a model of acute viral infection.

9. DeWitt WS III, Smith A, Schoch G, Hansen JA, Matsen FA IV, Bradley P. Elife. 2018 Aug 28;7. Human T cell receptor occurrence patterns encode immune history, genetic background, and receptor specificity.

10. Dines JN, Manley TJ, Svejnoha E, Simmons HM, Taniguchi R, Klinger M, Baldo L, Robins H. 2020. https://doi.org/10.1101/2020.08.17.20175158 medRxiv preprint. The ImmuneRACE Study: A Prospective Multicohort Study of Immune Response Action to COVID-19 Events with the ImmuneCODE Open Access Database.

11. Emerson R, DeWitt W, Vignali M, Gravley J, Hu J, Osborne E, Desmarais C, Klinger M, Carlson C, Hansen J, Rieder M, Robins H. 2017. Nature Genetics 49(5):659-655. Immunosequencing identifies signatures of cytomegalovirus exposure history and HLA-mediated effects on the T-cell repertoire.

12. Ferretti AP, Kula T, Wang Y, Nguyen DMV, Weinheimer A, Dunlap GS, Xu Q, Nabilsi N, Perullo CR, Cristofaro AW, Olivier KJ Jr, Baiamonte LB, Alistar AT, Whitman ED, Bertino SA, Chattopadhyay S, MacBeath G. 2020. https://doi.org/10.1101/2020.07.24.20161653 preprint. COVID-19 Patients Form Memory CD8+ T Cells that Recognize a Small Set of Shared Immunodominant Epitopes in SARS-CoV-2.

13. Glanville J, Huang H, Nau A, Hatton O, Wagar LE, Rubelt F, Ji X, Han A, Krams SM, Pettus C, Haas N, Arlehamn CSL, Sette A, Boyd SD, Scriba TJ, Martinez OM, Davis MM. 2017. Nature. Jul 6;547(7661):94-98. Identifying specificity groups in the T cell receptor repertoire.

14. Gonzalez-Galarza FF, McCabe A, Melo dos Santos EJ, Jones J, Takeshita L, Ortega-Rivera ND, Del Cid-Pavon GM, Ramsbottom K, Ghattaoraya G, Alfirevic A, Middleton D, Jones AR. 2020. Nucleic Acids Research 48:D1:D783-D788. Allele frequency net database (AFND) 2020 update: gold-standard data classification, open access genotype data and new query tools.

15. Grifoni A, Weiskopf D, Ramirez SI, Mateus J, Dan JM, Morderbacher CR, Rawlings SA, Sutherland A, Premkumar L, Jadi RS, Marrama D, de Silva AM, Frazier A, Carlin AF, Greenbaum JA, Peters B, Krammer F, Smith DM, Crotty S, Sette A. 2020. Cell 181(7):1489-1501. Targets of T Cell Responses to SARS-CoV-2 Coronavirus in Humans with COVID-19 Disease and Unexposed Individuals.

16. Klinger M, Pepin F, Wilkins J, Asbury T,Wittkop T, Zheng J, et al. 2015 PLoS ONE 10(10). Multiplex Identification of Antigen-Specific T Cell Receptors Using a Combination of Immune Assays and Immune Receptor Sequencing.

17. Long QX, Tang XJ, Shi QL, Li Q, Deng HJ, Yuan J, Hu JL, Xu W, Zhang Y, Lv FJ, Su K, Zhang F, Gong J, Wu B, Liu XM, Li JJ, Qiu JF, Chen J, Huang AL. 2020. Nature Medicine https://doi.org/10.1038/s41591-020-0965-6. Clinical and immunological assessment of asymptomatic SARS-CoV-2 infections.

18. Nelde A, Bilich T, Heitmann JS, Maringer Y, Salih HR, Roerden M, Lübke M, Bauer J, Rieth J, Wacker M, Peter A, Hörber S, Traenkle B, Kaiser PD, Rothbauer U, Becker M, Junker D, Krause G, Strengert M, Schneiderhan-Marra N, Templin MF, Joos TO, Kowalewski DJ, Stos-Zweifel V, Fehr M, Graf M, Gruber L-C, Rachfalski D, Preuß B, Hagelstein I, Märklin M, Bakchoul T, Gouttefangeas C, Kohlbacher O, Klein R, Stevanović S, Rammensee H-G, Walz JS. 2020: doi:10.21203/rs.3.rs-35331/v1 preprint. SARS-CoV-2 T-cell epitopes define heterologous and COVID-19-induced T-cell recognition.

19. Ng OW, Chia A, Tan AT, Jadi RS, Leong HN, Bertoletti A, Tan YJ. 2016. Vaccine 34 2008–2014. Memory T cell responses targeting the SARS coronavirus persist up to 11 years post-infection.

20. Nielsen M, Lundegaard C, Worning P, Lauemoller SL, Lamberth K, Buus S, Brunak S, Lund O. 2003. Protein Sci. 12:1007-17. Reliable prediction of T-cell epitopes using neural networks with novel sequence representations.

21. Nolan S, Vignali M, Klinger M, Dines J, Kaplan IM, Svejnoha E, Craft T, Boland K, Pesesky M, Gittelman R, Snyder TM, Gooley CJ, Semprini S, Cerchione C, Mazza M, Delmonte OM, Dobbs K, Carreño-Tarragona G, Barrio S, Martinelli G, Goldman J, Heath JR, Notarangelo LD, Carlson JM, Martinez-Lopez J, Robins H. 2020: doi: 10.21203/rs.3.rs-51964/v1 preprint. A large-scale database of T-cell receptor beta (TCRβ) sequences and binding associations from natural and synthetic exposure to SARS-CoV-2.

22. Peng H, Yang L-T, Wang L-Y, Li J, Huang J, Lu Z-Q, Koup RA, Bailer RT, Wu C-Y. 2006. Virology 351(2):466-475. Long-lived memory T lymphocyte responses against SARS coronavirus nucleocapsid protein in SARS-recovered patients.

23. Peng Y, Mentzer AJ, Liu G, Yao X, Yin Z, Dong D, Dejnirattisai W, Rostron T, Supasa P, Liu C, Lopez-Camacho C, Slon-campos J, Zhao Y, Stuart D, Paeson G, Grimes J, Antson F, Bayfield OW, Hawkins D, Ker DS, Turtle L, Subramaniam K, Thomson P, Zhang P, Dold C, Ratcliff J, Simmonds P, de Silva T, Sopp P, Wellington D, Rajapaksa U, Chen YL, Salio M, Napolitani G, Paes W, Borrow P, Kessler B, Fry JW, Schwabe NF, Semple MG, Baillie KJ, Moore S, Openshaw PJM, Ansari A, Dunachie S, Barnes E, Frater J, Kerr G, Goulder P, Lockett T, Levin R, Oxford Immunology Network Covid-19 Response T cell Consortium, Cornall RJ, Conlon C, Klenerman P, McMichael A, Screaton G, Mongkolsapaya J, Knight JC, Ogg G, Dong T. 2020 bioRxiv 2020.06.05.134551. Broad and strong memory CD4+ and CD8+ T cells induced by SARS-CoV-2 in UK convalescent COVID-19 patients.

24. Preussman W, Rytlewski J, Wilmott J, Mihm MC Jr, Attrill GH, Dyring-Andersen B, Fields P, Zhan Q, Colebatch AJ, Ferguson PM, Thompson JF, Kallenbach K, Yusko E, Clark RA, Robins H, Scoyler RA, Kupper TS. 2020. Nature Cancer 1:197-209. Molecular analysis of primary melanoma T cells identifies patients at risk for metastatic recurrence.

25. Robins HS, Campregher PV, Srivastava SK, Wacher A, Turtle CJ, Kahsai O, Riddell SR, Warren EH, Carlson CS. 2009. Blood 114(19):4099-4107. Comprehensive assessment of T-cell receptor β-chain diversity in αβ T cells.

26. Robins HS, Desmarais C, Matthis J, Livingston R, Andriesen J, Reijonen H, Nepom G, Yee C, Cerosaletti K. 2012. J. Immunol. Methods 375(1-2):14-9. Ultra-sensitive detection of rare T cell clones.

27. Robins, H. 2013. Curr Opin Immunol 25(5):646-52. Immunosequencing: applications of immune repertoire deep sequencing.

28. Sayers EW, Cavanaugh M, Clark K, Ostell J, Pruitt KD, Karsch-Mizrachi I. 2019. Nucleic Acids Research, 47(D1:D94-D99). GenBank.

29. Sekine T, Perez-Potti A, Rivera-Ballesteros O, Stralin K, Gorin JB, Olsson A, Llewellyn-Lacey S, Kamal H, Bogdanovic G, Muschiol S, Wullimann DJ, Kammann T, Emgard J, Parrot T, Folkesson E, Rooyackers O, Eriksson LI, Sonnerborg A, Allander T, Albert J, Nielsen M, Klingstrom J, Gredmark-Russ S, Bjorkstrom NK, Sandberg JK, Price DA, Ljunggren HG, Aleman S, Buggert M, Karolinska COVID-19 Study Group. 2020 bioRxiv 2020.06.29.174888. Robust T cell immunity in convalescent individuals with asymptomatic or mild COVID-19.

30. Tang F, Quan Y, Xin Z-T, Wrammert J, Ma M-J, Lv H, Wang T-B, Yang H, Richardus JH, Liu W, Cao W-C. 2011. J Immunol 186(12):7264-7268. Lack of Peripheral Memory B Cell Responses in Recovered Patients with Severe Acute Respiratory Syndrome: A Six-Year Follow-Up Study.

31. Vabret N, Britton GJ, Gruber C, Hegde S, Kim J, Kuksin M, Levantovsky R, Malle L, Moreira A, Park MD, Pia L, Risson E, Saffern M, Salomé B, Esai Selvan M, Spindler MP, Tan J, van der Heide V, Gregory JK, Alexandropoulos K, Bhardwaj N, Brown BD, Greenbaum B, Gümüş ZH, Homann D, Horowitz A, Kamphorst AO, Curotto de Lafaille MA, Mehandru S, Merad M, Samstein RM, and the Sinai Immunology Review Project. 2020. Immunology 52(6):910-941. Immunology of COVID-19: Current State of the Science.

32. Venturi V, Price DA, Douek DC, Davenport MP. 2008. Nat Rev Immunol. Mar;8(3):231-8. The molecular basis for public T-cell responses.

33. Weiskopf D, Schmitz KS, Raadsen MP, Grifoni A, Okba NMA, Endeman H, van den Akker JPC, Molenkamp R, Koopmans MPG, van Gorp ECM, Haagmans BL, de Swart RL, Sette A, & de Vries RD. 2020 Science immunology, 5(48), eabd2071. Phenotype and kinetics of SARS-CoV-2-specific T cells in COVID-19 patients with acute respiratory distress syndrome.

34. Zhao J, Alshukairi AN, Baharoon SA, Ahmed WA, Bokhari AA, Nehdi AM, Layqah LA, Alghamdi MG, Al Gethamy MM, Dada AM, Khalid I, Boujelal M, Al Johani SM, Vogel L, Subbarao K, Mangalam A, Wu C, Ten Eyck P, Perlman S, Zhao J. 2017. Sci. Immunol. 2(14); doi:10.1126/sciimmunol.aan5393. Recovery from the Middle East respiratory syndrome is associated with antibody and T cell responses.

